# Tracking Epigenetic Biomarkers of Health and Aging During the Initial Year of Gender-Affirming Hormone Therapy

**DOI:** 10.1101/2025.03.01.25323163

**Authors:** Khyobeni Mozhui, Beck A. Henriksen, Lauren A. Bell, Ingrid Bretherton, Ada S. Cheung, Boris Novakovic

**Affiliations:** Department of Preventive Medicine, College of Medicine, University of Tennessee Health Science Center, Memphis, TN, USA; Department of Genetics, Genomics and Informatics, College of Medicine, University of Tennessee Health Science Center, Memphis, TN, USA; Department of Religious Studies, and Gender and Sexualities Studies Program, and Institute for Health Equity and Community Justice, Rhodes College, Memphis, TN, USA; Department of Medicine (Austin Health), The University of Melbourne, Parkville, VIC 3052, Australia; Department of Endocrinology, Austin Health, Heidelberg, VIC 3084, Australia; Infection, Immunity and Global Health, Murdoch Children’s Research Institute and Department of Paediatrics, The University of Melbourne, Parkville, VIC 3052, Australia

## Abstract

Gender-affirming hormone therapy (GAHT) is a necessary treatment for many transgender people, and there is a critical need to further improve treatment experience and mitigate possible risks. Here we investigated whether DNA methylation (DNAm) biomarkers of health and aging are modified during the first year of GAHT and whether these vary by treatment type. Cohort consisted of 13 trans women and 13 trans men. Sampling occurred at baseline (pre-GAHT), and at 6- and 12-month follow-up. We tracked the longitudinal dynamics of three epigenetic clocks (Horvath, Hannum, PhenoAge), DNA methylation-based telomere length (DNAmTL), and DunedinPACE. At baseline, the Horvath and Hannum showed accelerated epigenetic aging, particularly pronounced among trans men, while the PhenoAge and DunedinPACE showed lower pace of aging in both groups. This discrepancy may reflect possible effects of minority stress in an otherwise healthy cohort. While GAHT did not affect the three clocks, DNAmTL and DunedinPACE showed treatment-specific patterns but with notable inter-individual variability in trajectories. Trans women had increased DunedinPACE (estimate = 0.057, p=0.002) and slight DNAmTL gains (estimate = 0.024, ns); trans men exhibited stable to slight decline in DunedinPACE (estimate = −0.013, ns), and reduction in DNAmTL (estimate = −0.057, p=0.037). The marked heterogeneity is indicative of an individualized response to treatment and highlights the potential value of incorporating such biomarkers in comprehensive health monitoring. Our findings emphasize the need for larger, long-term studies to optimize personalized strategies for gender-affirming healthcare.

## Introduction

Gender-affirming hormone therapy (GAHT) has been a medical treatment since the 1920s, with formal medical guidelines for transgender healthcare in place for many decades ^[1, 2]^. Despite this long history, the transgender and non-binary communities continue to face substantial health challenges stemming from societal discrimination, minority stress, and persistent barriers to healthcare ^[3–7]^. Individuals who identify as trans and non-binary present with significant diversity, and while not all trans people seek medical interventions to transition, for those who do, the treatment can alleviate symptoms of depression and distress caused by the gender dysphoria (i.e., the anguish arising from the incongruence between one’s gender identity and one’s physical characteristics) ^[8–11]^. Due to this, GAHT is generally deemed safe and beneficial, and in some cases, a necessary medical treatment ^[12]^.

As with any medical intervention, exogenous sex hormones presents both benefits and risks. For the majority cisgender population, several large-scale studies have tracked the long-term effects of hormone therapies (HT) ^[13–16]^. Valuable data collected from these have been instrumental in guiding risk management and in optimizing the dosing, timing, and route of administration ^[17–21]^. Consequently, a personalized approach to HT is feasibly within reach for cisgender individuals. In contrast, such advances remain limited for trans individuals, as trans and non-binary people have historically been excluded from all aspects of biomedical research. While recent studies have begun documenting the risks and benefits of exogeneous hormones unique to trans people, for the most part, our knowledge on the long-term effects remains limited ^[22–24]^. Similarly, data on the diversity of sex and gender have been traditionally excluded from “biospecimen research”, the foundation of modern genomic and multi-omics studies (i.e., epigenomics, transcriptomics, metabolomics, etc.) ^[25]^. Including minority and vulnerable populations in research is essential for delivering equitable health care, and it is only in recent years that there have been efforts to build participatory research studies that also engage trans and non-binary people ^[26–30]^.

A major advancement to the field came in 2022 when Shepherd et al. conducted the first longitudinal study of genome-wide DNA methylation (DNAm) in a balanced cohort of trans women (TW) and trans men (TM) receiving feminizing and masculinizing treatments, respectively.^[31]^ DNAm is a covalent chemical modification to the DNA, involving the addition of a -methyl group to cytosine residues (canonically, a cytosine adjacent to a guanine; hence the term CpG (de)methylation). The work by Shepherd et al. provided the first glimpse of how the epigenome is restructured during the first 12 months of gender-affirming feminizing, and masculinizing therapies. This has important relevance to health as DNAm plays fundamental roles in defining cellular identity and function, development, metabolism, immune function, and overall health ^[32]^.

Genome-wide DNAm is particularly valuable because it can be used to quantify various biomarkers of health and aging. These machine-learning based “epigenetic clocks” and related biomarkers such as the DunedinPACE are by no means a replacement for comprehensive functional and clinical assessments ^[33]^. Additionally, for many, the healing and relief to the dysphoria brought about by the hormonal treatment will far outweigh concerns about the long-term biological processes of aging. However, these biomarkers are powerful tools with potential application in personalized medicine, and have utility in monitoring health, evaluating therapeutic interventions, and in predicting disease risk and life expectancy ^[34–38]^. Furthermore, these DNAm readouts are associated with stress, social disparities, and experience of discrimination, and could offer insights into the complex interplay between social factors, personal experiences and discrimination, treatment effects, and health outcomes ^[39–41]^.

Here we leveraged the Shepherd et al. methylome data to compute four well-established biomarkers of health and aging: the Horvath, Hannum, PhenoAge, and DunedinPACE ^[38, 42–44]^. Given the roles of estrogen as a telomerase activator and testosterone in blood cell proliferation and telomere attrition, we also included the DNAm-based telomere length estimator (DNAmTL) ^[45–48]^. Our work revealed divergent patterns in DNAmTL and DunedinPACE between feminizing and masculinizing treatments, while also highlighting substantial heterogeneity in individual trajectories that suggests a highly individualized response to treatment.

## Methods

### Study Design and Participants

We performed a secondary analysis of genome-wide DNAm data originally reported in Shepherd et al. ^[31]^. Participants were recruited from endocrinology outpatient and primary care clinics specializing in transgender health in Melbourne, Australia. Study protocols and ethical oversight are documented in the original publication ^[31]^. All participants provided written informed consent for biospecimen research. In brief, the cohort consisted of 13 TW and 13 TM between the ages of 18 and 73. Feminizing hormones for the TW consisted of oral estradiol with doses ranging from 1–8 mg/day for n=5, and transdermal estradiol (25–200 mcg/day) for n=8. Most in the TW (n=11) received additional androgen blockers (12.5 mg/day cyproterone acetate for n=6; 100–200 mg/day spironolactone for n=3; 100 mg/day progesterone for n=2). Masculinizing hormones for the TM consisted of 1000 mg of intramuscular (IM) testosterone undecanoate administered every 12-weeks for n=7; 250 mg of fortnightly IM testosterone enanthate for n=1; 1.25–5 g/day of transdermal 1% testosterone gel for n=5. The present study does not have access to the individual-level treatment information.

### DNA methylation data and biomarker computation

Venous Blood was collected at baseline (before the initiation of HT), and at 6-, and 12-month follow-up. DNA was extracted from the buffy coat, and following standard quality checks, were processed for bisulfite conversion and assayed on version 1 of the Illumina Infinium MethylationEPIC beadchip (version 1). Detailed description of raw data processing and probe filters are in ^[31]^. The filtered set of beta-values was downloaded from NCBI Gene Expression Omnibus (GEO accession ID GSE176394) and directly used to compute the following DNAm models of aging: the Horvath and Hannum clocks as the representative first-generation models of chronological age^[42, 43]^; the PhenoAge clock, a second-generation predictor of health and lifespan ^[44]^; the DNAm based estimator of telomere length (DNAmTL) ^[45]^; and the more recent pace of aging DunedinPACE ^[38]^. The three clocks and DNAmTL were computed using the methylclock R package ^[49]^. DunedinPACE was computed using the R codes provided in the original publication’s GitHub repository. Cell proportion estimation was implemented using the estimateCellProp() function in the Enmix R package using the “FlowSorted.Blood.EPIC” reference data ^[50]^.

### Statistical analyses

We applied bivariate analyses for comparisons with chronological age, and t-test for cross-sectional analysis between the TW and TM groups. To track the longitudinal changes in measures of epigenetic aging, we applied linear mixed-effects regressions that fitted the visit time (6- and 12-month relative to baseline) as a fixed categorical variable, and participant as a random intercept (the R codes are provided as footnotes under Table 2) ^[51]^. Model 1 adjusted for chronological age as cofactor. Model 2 additionally included body mass index (BMI) and estimated proportion of blood cells (Bcell + CD4T + CD8T + Monocytes + Neutrophil + Natural Killer cells). The mixed effects analyses were done in the TW and TM groups separately. Statistical significance of the fixed effects were based on the Satterthwaite’s method that is implemented in the lmerTest R package ^[52]^. Analyses and graphing were done using R (v4.3.2) or JMP (Pro 18).

## Results

### Performance of Epigenetic Models at Baseline

The TM participants were all under age 30 at baseline (18.03–29.50 years; mean = 23.27 ± 3.31). The TW participants had greater age range (21.14–73.25 years; mean = 38.77± 19.77).

Both the Horvath and Hannum clocks predicted older epigenetic ages for most participants (**Fig 1a**, **1b**). We refer to the predicted ages as DNAmAge, and the chronological age as chroAge. Based on the absolute age-deviation (AAD; computed as DNAmAge minus chroAge), TM on average presented with >7 years (Horvath) and >2 years (Hannum) of accelerated aging (**Table 1**). Another measure of clock performance is the linear fit between chroAge and DNAmAge. Both the Horvath and Hannum estimates were highly correlated with chroAge among TW (r of 0.97). Given the narrow age-range, the correlations were significant but weaker for TM (0.63 for Horvath, 0.73 for Hannum).

**Fig 1.**
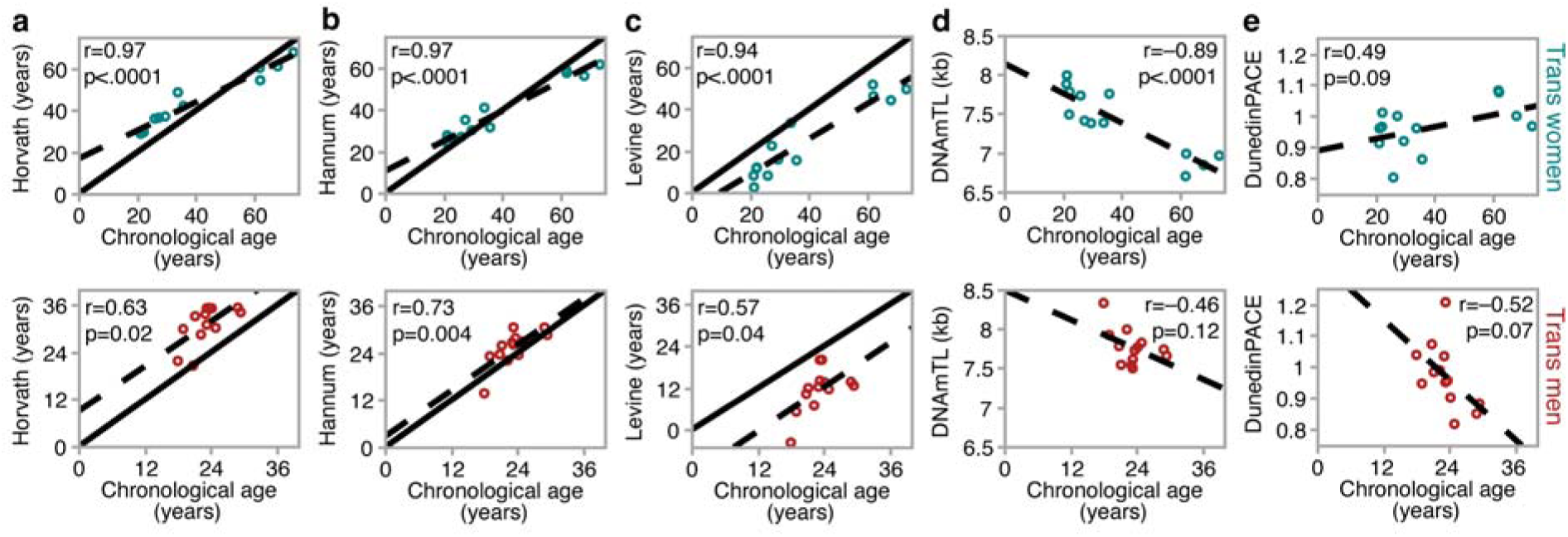
Chronological age versus the DNA methylation estimates. The scatter plots display how chronological age (chroAge) of participants relate to estimates computed by the **(a)** Horvath and **(b)** Hannum clocks, the **(c)** PhenoAge model of health and mortality, **(d)** the DNA methylation estimator of telomere length (DNAmTL), and **(e)** the DunedinPACE. These are baseline (pre-GAHT) values for the trans women (top; teal) and trans men (bottom; brick red). For the three models computed in units of years (**a–c**), the solid diagonal line indicates x=y (line where predicted DNAmAge perfectly matches chroAge); departure from this line is the “absolute age-deviation”. The dashed line corresponds to the linear regression fit.

**Table 1.** Measures of epigenetic aging and change over time.

| DNAm Model | Baseline | Feminizing hormone therapy |  |  |
| --- | --- | --- | --- | --- |
|  |  | 6-month | 12-month | 12-mo delta<sup>1</sup> |
| Horvath AAD (years) | 4.38±7.16 | 4.24±7.94 | 5.04±7.29 | 0.66 |
| Hannum AAD (years) | -0.16±6.46 | 0.21±6.27 | -0.47±6.2 | -0.31 |
| PhenoAge AAD (years) | -14.06±6.95 | -13.59±5.23 | -13.29±4.64 | 0.76 |
| DNAmTL (kb) | 7.41±0.42 | 7.36±0.35 | 7.43±0.42 | 0.02 |
| DunedinPACE | 0.964±0.078 | 1.007±0.084 | 0.998±0.088 | 0.034 |
| DNAm Model | Baseline | Masculinizing hormone therapy |  |  |
|  |  | 6-month | 12-month | 12-mo delta<sup>1</sup> |
| Horvath AAD (years) | 7.89±3.85 | 7.36±4.15 | 7.56±2.81 | -0.32 |
| Hannum AAD (years) | 2.09±2.99 | 2.19±3.06 | 2.16±2.45 | 0.07 |
| PhenoAge AAD (years) | -11.8±5.09 | -11.6±4.48 | -11.6±4.48 | -1.01 |
| DNAmTL (kb) | 7.77±0.23 | 7.73±0.2 | 7.7±0.16 | -0.07 |
| DunedinPACE | 0.972±0.103 | 0.966±0.079 | 0.952±0.083 | -0.020 |

<sup>1</sup>Computed as: (average at 12-month) – (average at baseline)

**Table 2.**
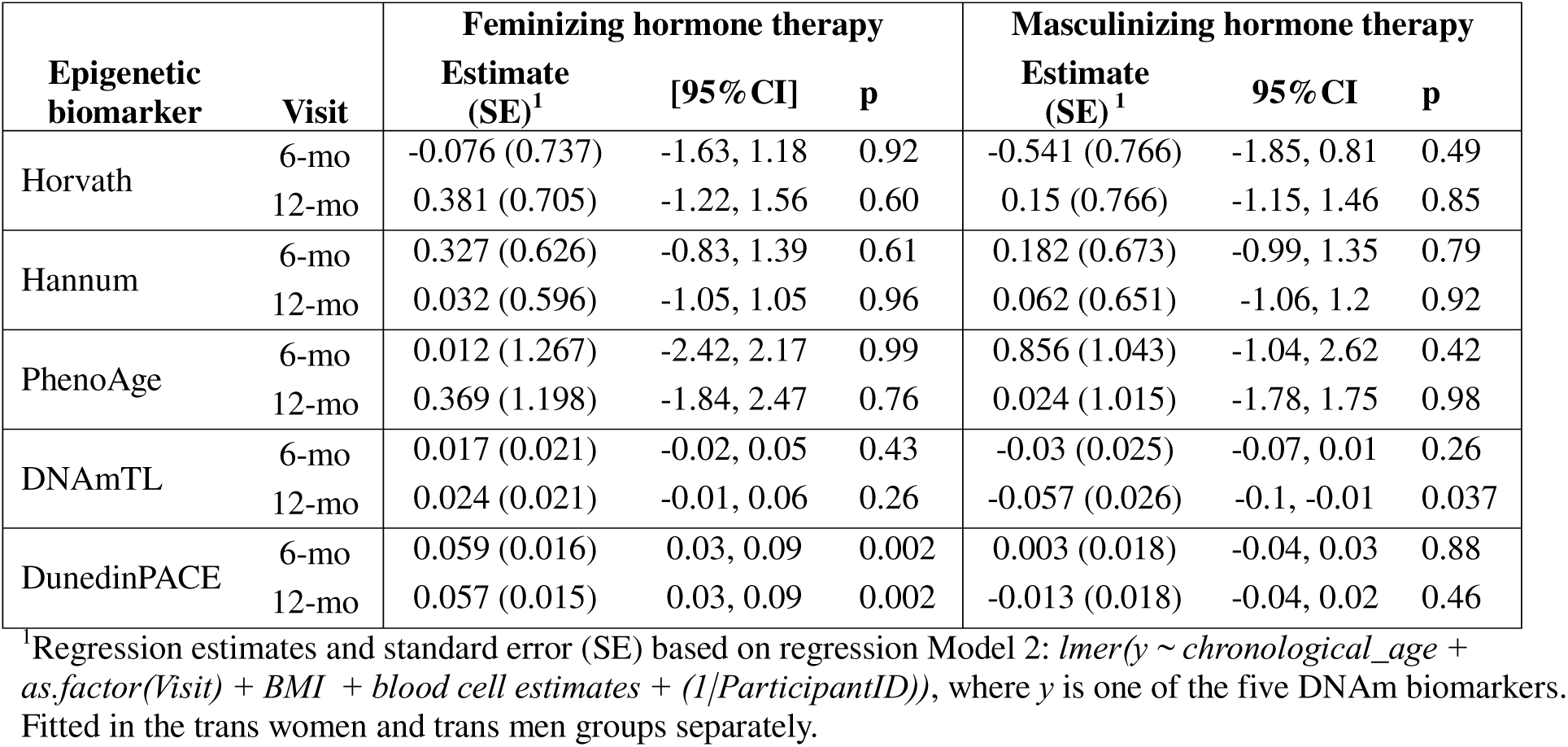
Results of linear mixed-effects analyses of the DNA methylation models at the two follow-up visits.

PhenoAge, although also computed in units of years, is not a chroAge predictor, and the computed number is more informative of physiological health and mortality risk ^[44, 53]^. PhenoAge showed the expected high deviation from chroAge, and in this cohort, it predicted younger epigenetic ages for both TW (–14 years) and TM (–12 years) (**Table 1**; **Fig 1c**). Correlation between chroAge and PhenoAge was strong for TW (r=0.93) but only modestly significant for TM (r=0.57).

DNAmTL is an indirect estimate of telomere length in kilobase (kb). It showed the expected inverse correlation with chroAge in both groups with older individuals having shorter predicted telomeres (**Fig 1d**). This inverse correlation was significant for TW due to the wide age range, and non-significant for the younger cohort of TM.

The DunedinPACE is expressed as a score with 1 indicating an average pace of aging, while <1 indicates slower, and >1 indicates faster pace of aging per year of chroAge.^[38]^ Similar to PhenoAge, DunedinPACE is strongly related to physiological and metabolic fitness. Suggesting a physiologically healthy cohort, DunedinPACE also showed lower pace of aging with mean scores of 0.964 for TW (∼roughly 3.6% slower pace of aging) and 0.972 for TM (∼2.8% slower pace of aging) (**Table 1**). Although the correlation did not reach statistical significance, the DunedinPACE had a positive r with chroAge among TW, and a negative r with chroAge among TM (**Fig 1e**).

### Longitudinal divergence in epigenetic aging

The mean change over the study period exhibited opposite trajectories between the masculinizing and feminizing therapies (**Table 1**). The Horvath, PhenoAge and DunedinPACE models had mean positive change among TW and mean negative change among TM. The DNAmTL also had contrasting changes with mean increase among TW, and mean loss among TM (**Table 1**).

Mixed-effects regression showed no significant modifications in age-deviation at the 6- or 12-month timepoints for the Horvath, Hannum, and PhenoAge (results for Model 2 in **Table 2**). DNAmTL showed slight but non-significant increases at both timepoints among TW. Among TM, there was a decrease in DNAmTL at 12-month relative to baseline that was modestly significant (estimate = −0.057, p=0.037; **Table 2**). This possible shortening of telomeres at the 12-month timepoint with masculinizing treatment was also detected by Model 1, albeit statistically weaker (estimate = –0.050; p = 0.18). The magnitudes of change at the two timepoints relative to baseline are shown in **Fig 2a**.

**Fig 2.**
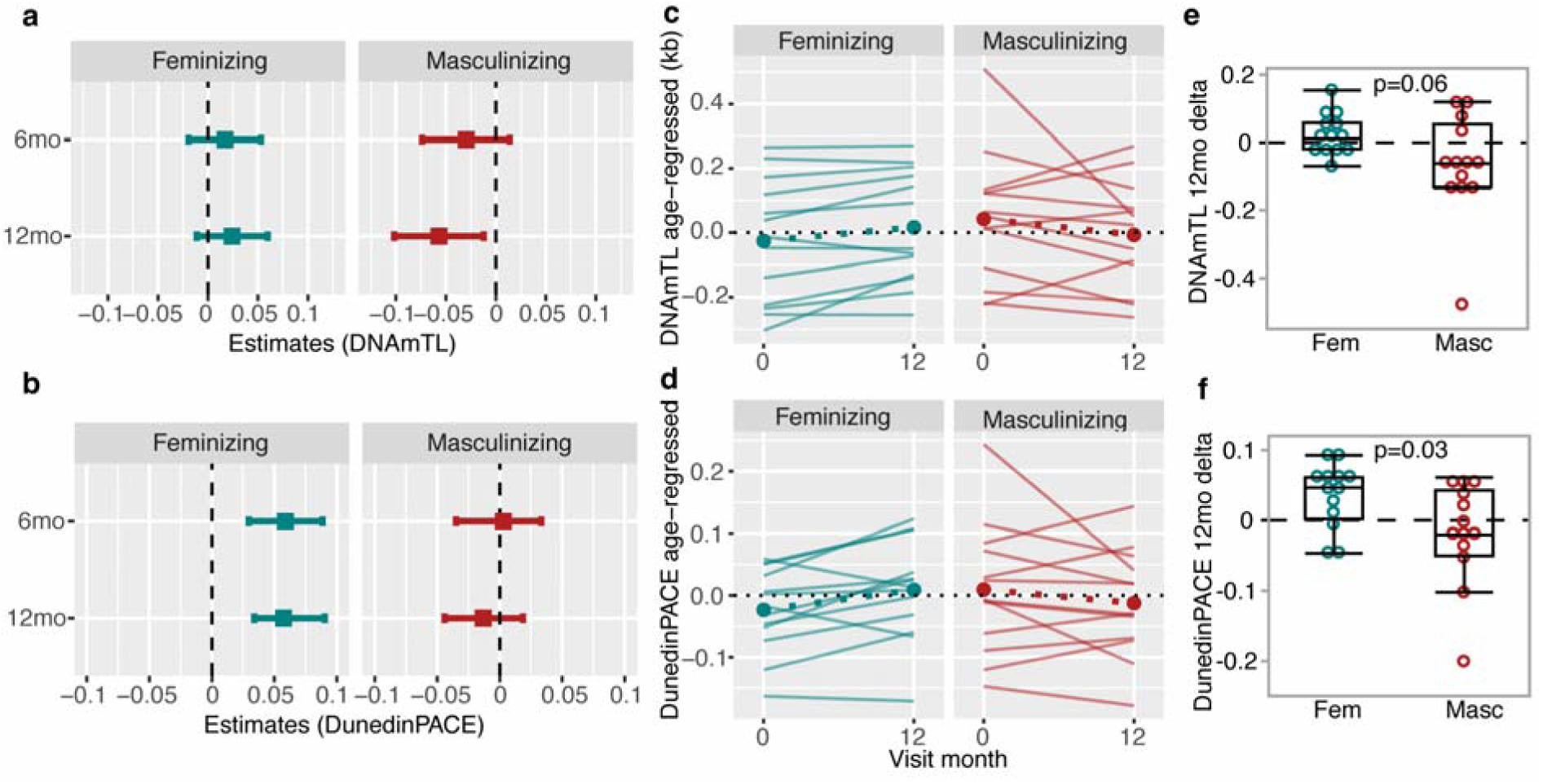
Longitudinal changes in DNAmTL and DunendinPACE. **(a)** Regression estimates (95% confidence interval) show the change in DNAmTL at the 6- and 12-month relative to baseline (teal denotes feminizing, and brick red denotes masculinizing hormone therapies). The model was adjusted for chronological age, BMI, and blood cell heterogeneity. **(b)** Similar full model regression estimates for DunedinPACE. The spaghetti plots show the age-adjusted individual changes in **(c)** DNAmTL, and **(d)** DunedinPACE from baseline to 12-month. To more clearly display the within-individual changes independent of age, the values are residuals after regression on chronological age. The dotted lines denote the average change. For each participant, change in **(e)** DNAmTL and **(f)** DunedinPACE was computed as unadjusted delta = (value at 12-month) minus (value at baseline). There is high variability, and participants in both treatment groups have negative (decline over time) as well as positive (gains over time) values. The p-values are the t-test based difference between the feminizing and masculinizing treatments.

DunedinPACE consistently increased in TW at both timepoints (**Fig 2b**; **Table 2**) and this increase over time was also detected by Model 1 for the 6- (estimate = 0.046; p = 0.002), and 12-month timepoints (estimate = 0.032; p = 0.02). For the TM group, DunedinPACE remained stable or decreased slightly by 12-month (**Table 2**; **Fig 2b**). Individual trajectories over the 12 months revealed substantial inter-individual heterogeneity (**Fig 2c**, **2d**; note that due to the varying levels of correlation with age and wide age range among TW, these longitudinal plots are age-regressed values). The 12-month change in DNAmTL in TM ranged from –0.478 kb to +0.118 kb (**Fig 2e**). Similarly, the 12-month change in DunedinPACE TW ranged from –0.047 to +0.093 (**Fig 2f**).

## Discussion

Our study provides initial evidence that there are measurable changes in DunedinPACE and DNAmTL during the first year of GAHT. The longitudinal slopes suggest divergence between feminizing and masculinizing treatments, though with notable inter-individual variability. However, the small sample size and the relatively short time span necessitate cautious interpretation. Furthermore, it is important to note that these machine learning models of epigenetic aging are influenced by the populations on which they are trained, and the present cohort represents a minority group that may have encountered greater life challenges than the general population ^[54]^.

The five selected DNAm based predictive models were developed to capture different facets of the complex processes of biological aging. The Horvath and Hannum are the first-generation “classical clocks” that were trained exclusively on chroAge and were designed to capture the generalized time-dependent patterns of aging ^[42, 43]^. Due to the small sample size and the study cohort, we employed the AAD method to estimate the rate of epigenetic aging (i.e., the simple difference between predicted versus known chroAge; this is also referred to as absolute age-acceleration) ^[55]^. The Hannum clock detected a slightly accelerated aging among TM but did not show significant longitudinal changes in either group. The Horvath clock also showed advanced epigenetic aging that was particularly pronounced in TM but remained stable over the study period. A potential explanation for the higher accelerated aging detected especially by the Horvath clock is the known eroding effect that negative social experiences have on the epigenome ^[56, 57]^. Previous studies have shown that epigenetic clocks, including the Horvath and Hannum, are accelerated by psychosocial stress, adversity, and discrimination ^[41, 58–61]^. Our observations align with the growing evidence that minority stress and healthcare barriers exert cumulative effects on the fundamental processes of aging ^[39, 62–65]^. However, as these clocks did not change significantly over the 12 months in both groups, we can infer that GAHT does not impact the overall aging process as estimated by these specific DNAm models.

The PhenoAge is conceptually slightly distinct as its training parameters placed emphasis on blood-based clinical measures (e.g., blood cell counts, glucose, creatinine, etc.), and the algorithm estimates the “phenotypic age” based on physiological traits that are correlated with aging ^[44, 66]^. While not used as an accurate predictor of chroAge, the relative age-deviation (aka, relative age-acceleration ^[55]^) derived from the PhenoAge outperforms the first-generation clocks in predicting future health outcomes and mortality risks ^[67, 68]^. Both the TW and TM showed mean negative AAD for PhenoAge at baseline that suggests a physiologically healthy cohort. The was also no significant longitudinal change in PhenoAge over the 12 months.

The DunedinPACE too was trained on blood-based clinical measures as well as several indicators of health and cardiovascular fitness (e.g., leptin, BMI, blood lipids, VO_2_Max), and this too estimated a slower pace of aging in both TW and TM. However, the most robust longitudinal change we detected was the consistent increase in DunedinPACE in TW, while TM showed either stable or a slight decrease. This observation merits careful consideration in the context of known cardiovascular and metabolic effects of estrogen therapy. This finding is particularly pertinent given that DunedinPACE was trained on longitudinal patterns in health-related parameters and is predictive of cardiovascular disease (CVD) and mortality risk ^[38, 69, 70]^. In postmenopausal women, despite the cardioprotective effect of estrogen ^[71–73]^, HT is associated with increased CVD risk ^[13, 17, 18]^. Correspondingly, recent observational and electronic health record studies have revealed an elevated risk of cardiovascular events and CVD-related mortality among TW on GAHT as compared to the general male and female populations ^[23, 24]^. For TM, risks for cardiovascular-specific events appear comparable to the general male population.

We must emphasize that we cannot determine causality from these observations. Instead, it raises several important questions. Do the changes in the endocrine environment and metabolic adaptation influence the DunedinPACE among TW? Might these effects relate to dosage, timing, administration route, or other adjuvant therapies? Alternatively, does the increase in pace of aging among TW primarily reflect psychosocial stressors during the initial phase of hormonal transition? Regarding the psychosocial domain, DunedinPACE is a remarkably sensitive biomarker of adversity, stress, and discrimination ^[39, 40, 58, 65]^. Recent cohort studies in the Netherlands and the UK revealed that TW and TM have disproportionally higher excess mortality from external causes including suicide and homicide compared to the general population ^[6, 24]^. And while each person’s transition experience is unique and it will be an oversimplification to make broad comparisons, there is data that indicates that TW experience higher levels of stigma due to greater visibility and prejudice ^[74]^, and perhaps due to changes in imposed gender roles and social hierarchy and status. Thus, the interplay of hormonal treatment effects with societal and interpersonal experiences presents a plausible alternate explanation for the differences in DunedinPACE trajectories between TW and TM.

The divergent trajectories in DNAmTL also raise questions about the differential effects of estrogen versus testosterone on biological processes. DNAmTL is a good estimator of telomere length but is not a direct measurement and may be more closely related to the proliferative history of cells ^[45]^. In the general population, females typically have longer telomeres in circulating blood than males, potentially contributing to the sexual dimorphism in mammalian aging and longevity ^[75]^. Our observations suggest a slight telomere increase in TW, and a decrease in TM over the 12 months. This is consistent with estrogen’s role as a telomerase activator, and HT among postmenopausal women may also be associated with longer telomeres.^[76]^ On the other hand, testosterone activates cell proliferation that can result in telomere attrition ^[46, 47]^.

However, the health implications of GAHT-associated DNAmTL changes remain unclear. This uncertainty also applies to the long-term significance of increased DunedinPACE among TW. While elevated pace of aging carries negative health implications, in the context of GAHT, there has been no research effort to systematically examine how such epigenetic biomarkers relate to long-term health and clinical outcomes. Furthermore, 12 months is a relatively short period, and we do not know whether the observed changes stabilize, progress, or even reverse over time.

Although the longitudinal sampling is a powerful design, the present work is limited by the small sample size and the heterogeneity in treatment regimens. Additionally, our study did not factor in differences in demographic and socioeconomic variables. Our findings strongly warrant the need for larger-scale, extended-duration follow-up studies that incorporate mental health and psychosocial variables. Long-term study of GAHT in trans populations offers dual benefits. First is the potential for improving and personalizing gender-affirming health care by leveraging innovative health and risk monitoring tools. Secondly, transgender people present a very relevant study paradigm to investigate the complex roles of sex hormones on fundamental biological mechanism that could lead to novel and generalizable insights on how sex and gender influence health and disease risks ^[29, 71, 77]^.

In conclusion, this preliminary work revealed accelerated epigenetic aging based on the Horvath clock that did not change over the first 12 months of GAHT. DunedinPACE changes suggested a potential contribution of feminizing treatment on accelerating cardiovascular risk, whereas changes in DNAmTL suggested decrease in telomere length with masculinizing hormone therapy. However, the observed heterogeneity in participant trajectories is indicative of the complex interplay between hormonal interventions and individual biology and experience. Taken together, our findings emphasize the need for close monitoring and a personalized approach to gender-affirming health care. Future large-scale research efforts that focus on patient characteristics, interpersonal and societal experiences, clinical measures and biomarkers will be critical in guiding treatment optimization that can lead to more effective strategies to maximize benefits of GAHT whilst minimizing potential risks.

## Acknowledgements

B Novakovic and A.S. Cheung are supported by the Australian National Health and Medical Research Council (NHMRC) Investigator Grants (1173314 and 2008956, respectively) and the Allen Distinguished Investigator program, a Paul G. Allen Frontiers Group advised program of the Paul G. Allen Family Foundation.

## Authors’ Contributions

K. Mozhui conceptualized the study, performed analysis, and contributed to drafting the original manuscript. B.A. Henriksen contributed to contextualizing the findings and drafting the original manuscript, and review and editing. L.A. Bell contributed to manuscript review and editing. I. Bretherton undertook clinical data and sample collection, and review. A.S. Cheung conceptualized and supervised the original clinical study, clinical data and sample collection, and contributed to review and editing. B. Novakovic conceptualized and supervised the original molecular study and bioinformatic analysis and contributed to manuscript review and editing. All authors approved the final manuscript.

## Data Availability

Deidentified data available on NCBI Gene Expression Omnibus. GEO accession ID GSE176394.

